# SURGERY VERSUS CONSERVATIVE STRATEGIES IN PATIENTS WITH ROTATOR CUFF TEAR OF THE SHOULDER: A SYSTEMATIC REVIEW WITH META-ANALYSIS

**DOI:** 10.1101/2020.07.13.20153015

**Authors:** Fabrizio Brindisino, Mattia Salomon, Silvia Giagio, Chiara Pastore, Tiziano Innocenti

**Affiliations:** Department of Medicine and Health Science “Vincenzo Tiberio”, University of Molise, Campobasso, Italy; Department of Clinical Science and Traslational Medicine, University of Roma “Tor Vergata”, Roma, Italy; Department of Biomedical and Neuromotor Science (DIBINEM), Alma Mater Studiorum, University of Bologna, Bologna, Italy; Centro Sanitario Riabilitativo FisicaMente - Predazzo (TN), Italy; Department of Neuroscience, Rehabilitation, Ophthalmology, Genetics, Maternal and Child Health, University of Genova, Campus of Savona, Italy

**Author notes:** **CORRESPONDING AUTHOR** Fabrizio Brindisino, via della Libertà, 14. 73023, Lizzanello (Le), ITALY, mail.

## Abstract

Shoulder pain (SP) is one of the most common musculoskeletal complaints 1 and it can negatively affect the correct movement of the upper limb, night rest, daily life activity, work and sports performances and autonomy 2-4.

Rotator cuff (RC) disease represents the most common cause of SP and it is responsible for up to 70% of all shoulder related visits to clinicians 5.

RC tears are generally considered to be a normal imaging result and a age related disorder 6, when we consider patients over 5th decade. Infact, RC tears are present between 20% to 54% of subjects aged between 60 and 80 years 7; moreover full-thickness RC tears can be evident in approximately 20% of patients over 65 years old 8.

RC tears have been widely studied and a lot of management strategies of patients with RC tears are actually available in literature 9,10; even if successful results have been achieved for both conservative and surgical treatment of RC tears, optimal management and best choice treatments for patient with RC tears are still unknown and debated 11,12.

Generally, conservative treatments were often administered in partial thickness RC tear, while surgery was judged as better option for massive tear 13. Furthermore, conservative treatment has often been advocated for older patients with comorbidities, while surgery is suggested for younger people 14,15. Lastly, physiotherapy did not reach structural healing of the tear, however successful rate was reported also after conservative treatment of massive tears: nevertheless, some concerns persist about the enlargement of the anatomical lesion and following loss of strength and pain persistence 7.

In the USA, in 2006, the annual incidence of surgery for RC tear was 98 procedures per 100,000 inhabitants and the incidence was increased form the application of the arthroscopic strategies 16,17, moreover, despite being considered as a successful treatment option, surgical treatment is estimated to cause from 20 to 90% rate of re-tear after surgery 18,19.

In such a framework of uncertainty on optimal management, several randomized controlled trials (RCTs) have been structured to compare the efficacy of surgical and conservative treatments for patient with any type of RC tears; results coming from different studies are often contradictory and substantially influenced by the recruited sample characteristics.

The aim of this systematic review is to analyze the results of randomized controlled trials which compare surgical and conservative treatments for patient with any type of RC tear through meta-analysis. Furthermore, this study also aims to know which are the most common indication to surgery: authors would like to understand if the presence of structural failure at the imaging assessment, the presence of pain refractory to conservative treatment, the presence of strength deficit or the combination of this mentioned elements are considered as decision criteria for choosing a surgery approach.

## Background

Shoulder pain (SP) is one of the most common musculoskeletal complaints ^1^ and it can negatively affect the correct movement of the upper limb, night rest, daily life activity, work and sports performances and autonomy ^2-4^.

Rotator cuff (RC) disease represents the most common cause of SP and it is responsible for up to 70% of all shoulder related visits to clinicians ^5^.

RC tears are generally considered to be a normal imaging result and a age related disorder ^6^, when we consider patients over 5th decade. Infact, RC tears are present between 20% to 54% of subjects aged between 60 and 80 years ^7^; moreover full-thickness RC tears can be evident in approximately 20% of patients over 65 years old ^8^.

RC tears have been widely studied and a lot of management strategies of patients with RC tears are actually available in literature ^9,10;^ even if successful results have been achieved for both conservative and surgical treatment of RC tears, optimal management and best choice treatments for patient with RC tears are still unknown and debated ^11,12^.

Generally, conservative treatments were often administered in partial thickness RC tear, while surgery was judged as better option for massive tear ^13^. Furthermore, conservative treatment has often been advocated for older patients with comorbidities, while surgery is suggested for younger people ^14,15^. Lastly, physiotherapy did not reach structural healing of the tear, however successful rate was reported also after conservative treatment of massive tears: nevertheless, some concerns persist about the enlargement of the anatomical lesion and following loss of strength and pain persistence ^7^.

In the USA, in 2006, the annual incidence of surgery for RC tear was 98 procedures per 100,000 inhabitants and the incidence was increased form the application of the arthroscopic strategies ^16,17,^ moreover, despite being considered as a successful treatment option, surgical treatment is estimated to cause from 20 to 90% rate of re-tear after surgery ^18,19^.

## METHODS

This systematic review will be conduct in accordance with the Preferred Reporting Items for Systematic Reviews and MetaAnalyses (PRISMA) statement ^20^. This review will be register on Prospero database.

The research will be conducted in 4 databases: MEDLINE, Cochrane Library (CENTRAL database), PEDro Database and Scopus, and the results will be included without date restriction until the 12^th^ of July 2020, considering studies published in English, Italian and German languages.

In addition, a manual search will be performed on the reference lists of included articles and other grey literature sources (eg. Google Scholar). References list of identified articles and reviews will be also checked for any relevancy.

### Inclusion and exclusion criteria

#### Participants/population

The inclusion criteria will be the following: adult (>18 years old) population (male and female) with any type of traumatic and atraumatic rotator cuff tear. while exclusion criteria will be: patients with shoulder fractures or shoulder dislocation (acute or recurrent), frozen shoulder (primary and secondary), any previous shoulder surgery procedure (regardless of shoulder pathology), tumors, infections, any systemic disease (e.g. rheumatoid arthritis).

#### Studies design selection

Only randomized controlled trial (RCTs) will be included.

#### Main outcomes to investigate

Main outcomes to be investigated will be Pain, Range of Motion (active and passive), function, disability, quality of life, muscle strength, patient (and clinician) satisfaction, willingness to avoid surgery. Furthermore, authors intend to investigate, when possible, additional outcomes such as return to work, return to sport, patient-perceived quality of treatment, adverse events, time for recovery.

#### Search strategy and Data extraction

Search strategy for each database is shown in table 1. Two review authors (F.B., M.S.) independently will select trials for possible inclusion according to inclusion/exclusion criteria.. Titles and abstracts will be screened and studies will be categorized into the following groups: a) possibly relevant trials that meet the inclusion criteria and trials from which it was not possible to determine whether they met the criteria either from their title or abstract; b) studies that did not meet the inclusion criteria and therefore that will be excluded.

If a title or abstract will suggest that the trial could be eligible for inclusion or if there will be inconsistent elements to clarify whether or not to include it, authors will obtain a full-text version of the article. Two review authors will then independently assess it to determine whether it met the inclusion criteria.

To sort out the included studies and extract data, a standardized planned Excel form will be used.

The extraction form will be filled in by two authors (S.G and C.P) alternately, with mutual check on each entry. Disagreements between reviewers will be solved resolved by the third reviewer assessment of the article (T.I.)

The following data will be extracted: patient’s characteristics, selection criteria, type of interventions, follow-up periods, outcome measures and results.

When additional data will be required, we will contact the trial’s authors. We will report in the extraction form Data that can be imputed or calculated (e.g. standard deviations calculated from standard errors, P values, confidence intervals, imputed from graphs, from standard deviations in other trials). Any disagreements and issues will be resolved by the consultation with a third author (T.I.).

To prevent selective inclusion of data based on the results, we will use the following a priori defined decision rules to select data from trials: a) Where trialists reported both final values and change from baseline values for the same outcome, we will extract final values; b) Where trialists reported both unadjusted and adjusted values for the same outcome, we will extract unadjusted values; c) Where trialists reported data analysed based on the intention-to-treat (ITT) sample and another sample (e.g. per-protocol, as-treated), we will extract ITT-analysed data.

#### Risk of bias (quality) assessment

The risk of bias of individual studies assessment will be considered for either the outcome and the level of evidence. Two authors will independently assess the risk of bias in included studies by using the Revised Cochrane risk-of-bias tool for randomized trials (RoB 2). RoB graph will be created through RobVis visualization tool ^21^.

Any disagreement over the assessment of risk of bias studies will be resolved through discussion between all authors.

#### Strategy for data synthesis

Meta-analysis of data will be conducted. Statistical heterogeneity of studies will be evaluated using the I^2^-test. An I^2^ value greater than 60% will be considered as substantial heterogeneity. The random effects model will be used as a conservative approach to account for different sources of variation among studies. The significance level will be defined as P<0.05. All the P values will be two-sided, while the weight of studies will be calculated according to the reciprocal of their variance. In our study, we will synthesize the data using a random-effect method, with risk ratios (RR) as effect size for dichotomous outcomes and standardized mean differences (SMD) for continuous outcomes.

If a substantial heterogeneity will be found, a sensitivity analysis according to various factors, such as study populations and study quality to investigate source of heterogeneity will be conducted. We will also assess evidence of publication bias with funnel plots methods.

Where multiple trial arms are reported in a single trial, we will include only the relevant arms. For studies containing more than two intervention groups, making multiple pair-wise comparisons between all possible pairs of intervention groups possible, we will include the same group of participants only once in the meta-analysis.

All analyses will be performed using Review Manager Version 5.3 software (http://www.cochrane.org) for statistical analysis.

#### Analysis of subgroups or subsets

Because of conservative treatments are different each other, we will compare surgery with different conservative options, as detailed below:

- Surgery versus Manual Therapy (Manual techniques + exercise therapy + stretching) for traumatic tears
- Surgery versus Manual Therapy (Manual techniques + exercise therapy + stretching) for atraumatic tears
- Surgery versus physical agents
- Surgery versus drug (OS)
- Surgery versus injection
- Surgery with additional procedures (e.g. LHBT tenotomy or tenodesis) versus conservative treatment
- Surgery without additional procedures (e.g. LHBT tenotomy or tenodesis) versus conservative treatment

In case of other further comparisons between surgery and conservative treatments from the included studies, authors will provide to extend subgroups analysis

#### Missing data

When required, we will contact trial authors to obtain data that were missing from the trial reports. For continuous outcomes, we will calculate the weight of the trial using the number of patients analysed at that time point. If the number of patients analysed will not present for each time point, we will use the number of randomised patients in each group at baseline. For dichotomous outcomes, we will use the final data for the events reported in each trial.

For continuous outcomes with no standard deviation (SD) reported, we will calculate SDs from standard errors (SEs), 95% confidence intervals (CIs) or P values. We will plan to impute SDs when we could not obtain any measurement of variance from the trial reports or by contacting the authors.

## RELEVANCE AND DISSEMINATION

We believe that results in this systematic review will increase clinicians knowledge about implications of surgical and conservative treatment in patients with rotator cuff tear. Furthermore this study will inform about the best treatment choice regarding different rotator cuff tears treatment options. Moreover this study will add relevant information that may contribute to improve the management of the patient with this kind of diseases.

The results of this research will be published in a peer-review journal and will be presented at relevant national and international scientific events.

## Data Availability

none

